# Detecting in-school transmission of SARS-CoV-2 from case ratios and documented clusters

**DOI:** 10.1101/2021.04.26.21256136

**Authors:** Kaitlyn E. Johnson, Michael Lachmann, Madison Stoddard, Remy Pasco, Spencer J. Fox, Lauren Ancel Meyers, Arijit Chakravarty

**Affiliations:** Department of Integrative Biology, University of Texas at Austin, Austin, TX; Santa Fe Institute, Santa Fe, NM; Fractal Therapeutics, Cambridge, MA, USA

**Keywords:** SARS-CoV-2, school transmission, K-12 education

## Abstract

Claims that in-person schooling has not amplified SARS-CoV-2 transmission are based on similar infection rates in schools and their surrounding communities and limited numbers of documented in-school transmission events. Simulations assuming high in-school transmission suggest that these metrics cannot exclude the possibility that transmission in schools exacerbated overall pandemic risks.

## Introduction

A recent CDC Commentary states there is “little evidence that schools have contributed meaningfully to transmission” [1], based on reports of similar infection rates in schools compared to the surrounding community [2,3] and the relative infrequency of documented school-based outbreaks [2–7]. However, infection rates in schools have been estimated primarily from testing of symptomatic students rather than proactive testing of asymptomatic students, which can lead to underestimation of the total attack rate since asymptomatic spread accounts for ∼60% of SARS-CoV-2 transmission [8], the majority of transmission comes from a minority of cases (overdispersion) [9], and children are more likely to experience asymptomatic infections than adults [10]. Here, we use mathematical models incorporating these transmission characteristics of SARS-CoV-2 to determine whether the ratio of infection rates in children versus adults coupled with the lack of detected in-school outbreaks is sufficient to rule out the possibility that schools are amplifying COVID-19 transmission.

## The Study

First, we assessed whether transmission risks in schools can be inferred based on the ratio of SARS-CoV-2 attack rates among school children versus the surrounding community. We use a Susceptible-Exposed-Infectious-Recovered (SEIR) compartmental model of SARS-CoV-2 transmission with age-specific contact rates [11] and location-specific transmission probabilities (Technical Appendix, Figure S1). The model assumes that all symptomatic infections are detected, with children having a lower probability of symptom presentation (21%) than adults (70%) [10]. We simulated two scenarios, one in which schools amplify transmission (*high risk*) and another in which they do not (*low risk*). The low risk scenario assumes a population-wide reproduction number of *R*_0_ = 1.1; the high risk scenario sets the transmission risk among children to correspond to an elevated in-school reproduction number of *R*_0_ = 2.5 (see Technical Appendix for details).

Under the high risk scenario, our model predicts a high SARS-CoV-2 attack rate among both school children and adults in the surrounding community, with school children having a higher infection rate (Figure 1A) but similar case detection rate (Figure 1B) compared to adults. Because children are much more likely to experience asymptomatic infections that go undetected [10], resulting in much lower reported to true case rates in children than adults [12], the ratio of reported cases in children versus adults is expected to drop well below one, even when the true incidence is much higher in children (Figure 1C). For comparison, a recent study in Wood County, Wisconsin [2] found that the ratio of reported cases among children attending in-person school versus all other members of the community was 0.57 (2,728 per 100,000 in children vs 4,785 in adults) by the 13th week of the school year (Figure 1C, horizontal line). By the 13th week of the simulation, the observed proportion is equally if not more consistent with the high risk scenario, suggesting that a low ratio of infection rates in school versus the community does not exclude the possibility that schools amplify community transmission of SARS-CoV-2 [10].

**Figure 1.**
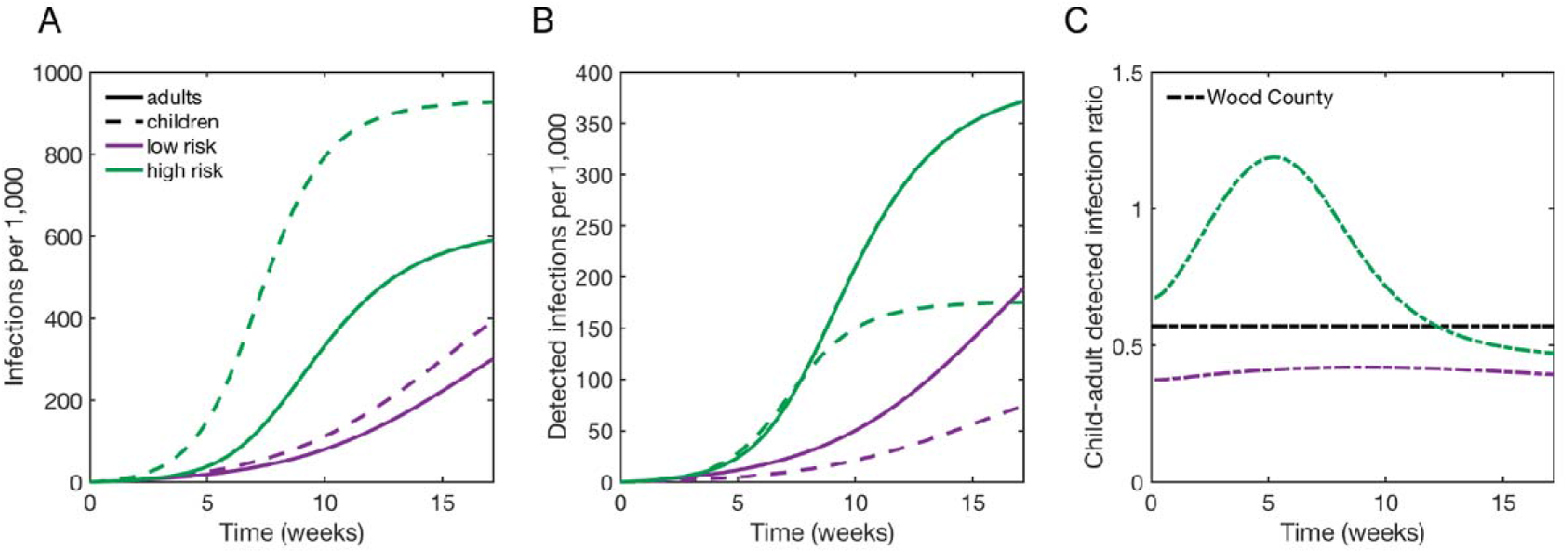
Projected infection and case detection rates under two different school scenarios. (A) Cumulative attack rates and (B) detected cases in children (dashed lines) and adults (solid lines) assuming that transmission rates in schools are either equal to the surrounding community (low risk, purple) or elevated (high risk, green). (C) The ratio of cumulative detected cases in children versus adults, under the same two scenarios. The black line indicates the ratio of reported cases in children attending in-person school (133 cases among 4,876 children) versus reported cases in adults (3,260 cases among ∼68,124 adults) in the surrounding community of Wood County, Wisconsin [2] after 13 weeks of in-school instruction. The projections assume a population-wide reproduction number of *R*_*0*_ = 1.1 in the low risk scenario and elevated in-school reproduction number of *R*_*0*_ = 2.5 in the high risk scenario, that children and adults have distinct symptomatic proportions [10], and that all symptomatic infections are detected.

Second, we estimated the likelihood of detecting outbreaks that occur in schools via *symptom-gated forward contact tracing*, which has been commonly used by schools in the US [2]. Detecting child-to-child transmission using this approach relies on the appearance and reporting of two consecutive symptomatic cases, connected by a transmission event. If we assume that only symptomatic cases are detected, that 21% of pediatric COVID-19 infections are symptomatic [10], and that asymptomatic and symptomatic infections are equally infectious [13], then roughly 4.4% of all child-to-child transmission events in schools would be detectable. Wood County [2] identified seven in-school transmission events via symptom-gated forward contact tracing, suggesting that there may have been over 150 child-to-child infections in the same time period. Using a Markov model of SARS-CoV-2 transmission that accounts for overdispersion in secondary cases [9] (Figure 2A, Technical Appendix, Figure S2), we estimate the chance that a detected symptomatic case will infect at least one child who also develops symptoms. As a result of overdispersion, most of these index cases are not expected to transmit to others [9]; of those that do, only a small fraction of the resulting secondary cases will be symptomatic [10]. Assuming a negative binomial distribution of secondary cases with mean (*R*_*0*_) of 2.5 and an overdispersion parameter *k* of 0.1 [9] (corresponding to the high risk in-school transmission scenario), we expect that only 28% of index cases would infect at least one child and only 16.5% [11.5%-22.0%] would infect at least one child that goes on to develop symptoms (Figure 2B). Given the difficulty of ascertaining child-to-child SARS-CoV-2 infections via symptom-gated forward contact tracing, low numbers of documented events do not necessarily indicate a lack of in-school transmission.

**Figure 2.**
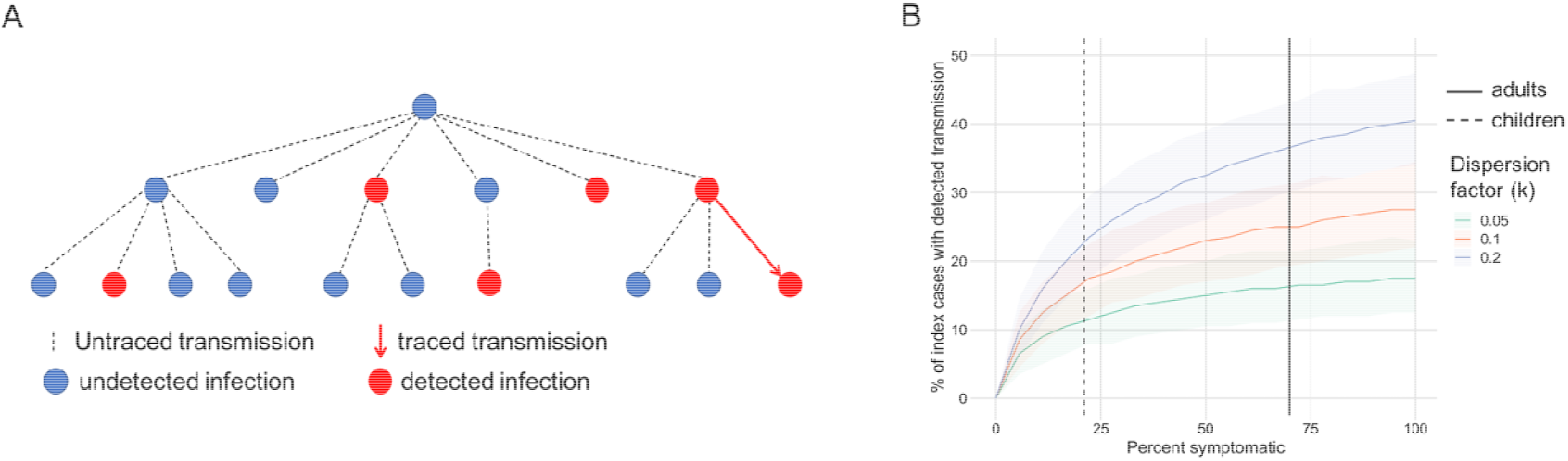
Detecting in-school transmission by symptom-gated forward contact tracing. A. Schematic illustration of a SARS-CoV-2 transmission tree demonstrating the impact of overdispersion and asymptomatic infections on the effectiveness of symptom-gated forward contact tracing. In this example, only 6 out of 17 infections and 1 out of 16 transmission events are detected. B. We estimate the percent of symptomatic index cases expected to have one or more detected secondary infections, under three different levels of overdispersion in the distribution of secondary infections, corresponding to the estimated median (*k=*0.1) and 95% CI (0.05-0.2) for SARS-CoV-2 [9]. As the dispersion factor (*k*) increases, the proportion of cases that cause at least one secondary infection also increases and the likelihood of detecting transmission events becomes more sensitive to the underlying symptomatic proportion. Dashed lines indicate estimated symptomatic proportions for children (21%) and adults (70%) [10]. Model simulations are based on 200 index cases and 1,000 repeated simulations.

Our estimates optimistically assume that all symptomatic cases are reported and all transmission events between successive symptomatic cases are detected. However, recent contact-tracing studies in the United States suggest that many named contacts are not successfully traced [14,15] and not all symptomatic contacts are willing to undergo testing [7], both of which would further undermine the detection of cases and transmission events in schools. On the other hand, we assume that only symptomatic infections are detected. Our findings do not pertain to schools that have established proactive (asymptomatic) surveillance testing programs [12,16] and contact-tracing programs that offer testing to all contacts, regardless of symptom status. Of note, in the case where an accurate picture of the true infection rates is possible, we found that as contacts between children and adults increased, total infection rates in the population rose, while the gap between infection rates in children and adults narrowed (Figure S3). As a final caveat, our model does not explicitly account for adults working in schools or children who are not in school; both of these groups would be expected to have infection risks associated with their environment, but symptomatic rates associated with their age group.

## Conclusions

In this work, we examined whether the absence of evidence of in-school transmission could be taken as evidence of absence. We simulated a scenario in which schools have a higher rate of SARS-CoV-2 transmission than the surrounding community and compared it to a scenario where the rate of transmission was similar between schools and the community. We found that the ratio of cases occurring in and out of schools and low numbers of documented in-school transmission events are not sufficient to distinguish these two scenarios. While there are compelling arguments for returning to in-person learning, particularly as SARS-CoV-2 vaccine coverage increases, a robust understanding of the limitations of the metrics being used to assess SARS-CoV-2 transmission is critical for risk management.

## Data Availability

All data referred to is published data from a recent study: https://www.cdc.gov/mmwr/volumes/70/wr/mm7004e3.htm

## Acknowledgments

We acknowledge the financial support from NIH Grant R01 AI151176 and CDC Grant U01IP001136. M.S. received funding from Fractal Therapeutics for the duration of this project. The funding organization did not play a role in the study design, data analysis, decision to publish or preparation of the manuscript.

## References

1. Honein MA, Barrios LC, Brooks JT. Data and Policy to Guide Opening Schools Safely to Limit the Spread of SARS-CoV-2 Infection. JAMA. 2021;325: 823–824.

2. Falk A, Benda A, Falk P, Steffen S, Wallace Z, Høeg TB. COVID-19 Cases and Transmission in 17 K-12 Schools - Wood County, Wisconsin, August 31-November 29, 2020. MMWR Morb Mortal Wkly Rep. 2021;70: 136–140.

3. Gandini S, Rainisio M, Iannuzzo ML, Bellerba F, Cecconi F, Scorrano L. No evidence of association between schools and SARS-CoV-2 second wave in Italy. doi:10.1101/2020.12.16.20248134

4. Zimmerman KO, Akinboyo IC, Brookhart MA, Boutzoukas AE, McGann K, Smith MJ, et al. Incidence and Secondary Transmission of SARS-CoV-2 Infections in Schools. Pediatrics. 2021. doi:10.1542/peds.2020-048090

5. Yung CF, Kam K-Q, Nadua KD, Chong CY, Tan NWH, Li J, et al. Novel Coronavirus 2019 Transmission Risk in Educational Settings. Clin Infect Dis. 2021;72: 1055–1058.

6. Macartney K, Quinn HE, Pillsbury AJ, Koirala A, Deng L, Winkler N, et al. Transmission of SARS-CoV-2 in Australian educational settings: a prospective cohort study. Lancet Child Adolesc Health. 2020;4: 807–816.

7. Doyle T, Kendrick K, Troelstrup T, Gumke M, Edwards J, Chapman S, et al. COVID-19 in Primary and Secondary School Settings During the First Semester of School Reopening - Florida, August-December 2020. MMWR Morb Mortal Wkly Rep. 2021;70: 437–441.

8. Johansson MA, Quandelacy TM, Kada S, Prasad PV, Steele M, Brooks JT, et al. SARS-CoV-2 Transmission From People Without COVID-19 Symptoms. JAMA Netw Open. 2021;4: e2035057.

9. Endo A, Centre for the Mathematical Modelling of Infectious Diseases COVID-19 Working Group, Abbott S, Kucharski AJ, Funk S. Estimating the overdispersion in COVID-19 transmission using outbreak sizes outside China. Wellcome Open Res. 2020;5: 67.

10. Davies NG, Klepac P, Liu Y, Prem K, Jit M, CMMID COVID-19 working group, et al. Age-dependent effects in the transmission and control of COVID-19 epidemics. Nat Med. 2020;26: 1205–1211.

11. Prem K, Cook AR, Jit M. Projecting social contact matrices in 152 countries using contact surveys and demographic data. PLoS Comput Biol. 2017;13: e1005697.

12. Crowe J, Schnaubelt AT, Schmidt-Bonne S, Angell K, Bai J, Eske T, et al. Pilot program for test-based SARS-CoV-2 screening and environmental monitoring in an urban public school district. bioRxiv. medRxiv; 2021. doi:10.1101/2021.04.14.21255036

13. Lee S, Kim T, Lee E, Lee C, Kim H, Rhee H, et al. Clinical Course and Molecular Viral Shedding Among Asymptomatic and Symptomatic Patients With SARS-CoV-2 Infection in a Community Treatment Center in the Republic of Korea. JAMA Intern Med. 2020. doi:10.1001/jamainternmed.2020.3862

14. Nadeem R. The Challenges of Contact Tracing as U.S. Battles COVID-19. 30 Oct 2020 [cited 15 Apr 2021]. Available: https://www.pewresearch.org/internet/2020/10/30/the-challenges-of-contact-tracing-as-u-s-battles-covid-19/

15. Hendrix MJ, Walde C, Findley K, Trotman R. Absence of Apparent Transmission of SARS-CoV-2 from Two Stylists After Exposure at a Hair Salon with a Universal Face Covering Policy - Springfield, Missouri, May 2020. MMWR Morb Mortal Wkly Rep. 2020;69: 930– 932.

16. Gillespie DL, Meyers LA, Lachmann M, Redd SC, Zenilman JM. The Experience of 2 Independent Schools With In-Person Learning During the COVID-19 Pandemic. J Sch Health. 2021;91: 347–355.

