## Appendix for "Detecting in-school transmission of SARS-CoV-2 from case ratios and documented clusters"

### Technical Appendix for "Detecting in-school transmission of SARS-CoV-2 from case ratios and documented clusters"

#### Two subpopulation SEIR model

The two-subpopulation SEIR model structure is diagrammed in Figure S1 and described in the equations below.

The model contains two compartments, children (assumed here to be synonymous with schools) and adults (assumed here to be synonymous with the community). While real-world school populations contain adult school staff, they comprise 5-10% of a typical school population and have been neglected here as a first approximation. As numerous studies have reported adult-to-adult transmission in the in-school setting [1–4], the adults working in schools can be considered in our model to be part of the "community". The issue of whether adults working in schools are at a higher risk of infection is beyond the scope of this work. Real-world populations also contain a small percentage of children who are not of school age, but these children would either be at home (and hence part of the community) or in daycares (which can be considered as functionally equivalent to schools). Thus, in the interests of parsimony, we have represented the four-compartment structure (children in schools, adults in schools, adults in the community, children in the community) as a two-compartment structure here, focusing on the interaction between children in schools and adults in the community.

The model assumes that the school-age children (or kids, $K$) and adults ($A$) have the ability to infect susceptibles both within their subpopulation and to some degree in the other subpopulation. To incorporate school transmission, we assume that all contacts between school-age children (denoted $\mu_{K<-K}$ for child-to-child) occur in the school setting and have a probability, $p_s$, of infection given contact with an infected child. All other contacts (between adults $\mu_{A<-A}$, from children to adults $\mu_{A<-K}$, from adults to children $\mu_{K<-A}$) are assumed to occur in the "community" and have a probability, $p_c$, of infection given contact with an infected individual. If schools are open and schools have higher transmission rates than communities, then $p_S > p_c$. If schools are closed, we set $p_S = p_c$, thereby assuming that children to children contacts occur in a risk-mitigated setting equivalent to the risk of other community interactions. The number of contacts within and between children and adults as described above were set for the baseline scenario based on the total POLYMOD matrices estimated for the United States [5]. Individuals transition between the states: susceptible (S), exposed (E), infected (I), and recovered (R). The K and A subscripts refer to the individual's status as a school-aged child (K) or an adult (A). The symbols $S_A$, $S_K$, $E_A$, $E_K$, $I_A$, $I_K$, $R_A$, and $R_K$ denote the number of people in that state. The model equations are given by:

$$\frac{dS_K}{dt} = -S_K(\frac{p_S\mu_{K<-K}I_K}{N_K} + \frac{p_C\mu_{K<-A}I_A}{N_A})$$
$$\frac{dE_K}{dt} = S_K(\frac{p_S\mu_{K<-K}I_K}{N_K} + \frac{p_C\mu_{K<-A}I_A}{N_A}) - \alpha E_K$$
$$\frac{dI_K}{dt} = \alpha E_K - \gamma I_K$$
$$\frac{dR_K}{dt} = \gamma I_K$$

$$\frac{dS_A}{dt} = -S_A(\frac{p_C\mu_{A<-A}I_A}{N_A} + \frac{p_C\mu_{A<-K}I_K}{N_K})$$
$$\frac{dE_A}{dt} = S_A(\frac{p_C\mu_{A<-A}I_A}{N_A} + \frac{p_C\mu_{A<-K}I_K}{N_K}) - \alpha E_A$$
$$\frac{dI_A}{dt} = \alpha E_A - \gamma I_A$$
$$\frac{dR_A}{dt} = \gamma I_A$$

Where $N_A$ and $N_K$ are the number of adults and children in the community respectively, $\alpha$ is the exposed rate, and $\gamma$ is the recovery rate.

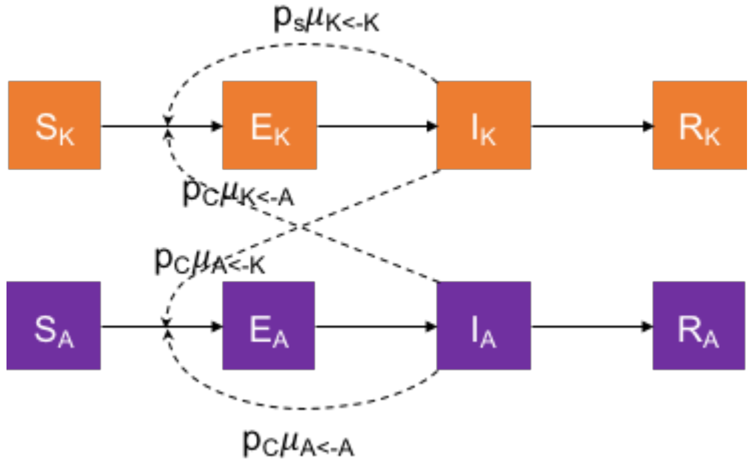


**Figure S1. Diagram of the two-population COVID-19 transmission model.** Upon exposure from either an infected adult or child, susceptible ($S_A$ or $S_K$) individuals progress to exposed ($E_A$ or $E_K$), from which they move to infected ($I_A$ or $I_K$), and eventually recover ($R_A$ or $R_K$). These individuals are no longer susceptible. The $p_i\mu_{j<-k}$ terms dictates the rate at which infected individuals from one subpopulation, *k*, infect another subpopulation, *j*, in a certain environment *i*.

The probability of transmission in the school and community are calculated based on the desired basic reproductive number, $R_{0i}$, within a group:

$$p_i = \frac{R_{0,i<-i}\gamma}{\mu_{i<-i}}$$

Detected cases in both adults and school-aged children were assumed to be symptom-based only, as the cited studies report that no surveillance screening was performed in schools and communities [6,7], and were simply a multiple of the number of true infections within each subpopulation.

$$cases = p_{sym} I$$

Model parameters and initial conditions are presented in Table S1. Parameters for the two scenarios displayed in Figure 1 are in Table S2.

Table S1: Model Parameters

| **Parameters** | **Value** |
|---|---|
| $\alpha$: transition rate from exposed to infectious | ⅓ [8] |
| $\gamma$: recovery rate | 1/10 [9] |
| $R_{0,S}$: school reproduction number | 2.5 |
| $R_{0,C}$: community reproduction number | 1.1 |
| $p_c$: probability of transmission in community | 0.0122 |
| $p_S$: probability of transmission in school | 0.0258 |
| $p_{sym,S}$: symptomatic proportion in school-age children | 0.21 [10] |
| $p_{sym,C}$: symptomatic proportion in community | 0.70 [10] |
| Initial number infected in the school | 3 per 1000 |
| Initial number infected in the community | 3 per 1000 |
| $\mu_{K<-A}$: number of contacts children have with adults | 5.3 [5] |
| $\mu_{A<-K}$: number of contacts adults have with children | 2.5 [5] |
| $\mu_{K<-K}$: number of contacts children have with kids | 9.7 [5] |
| $\mu_{A<-A}$: number of contacts adults have with children | 9.0 [5] |

Table S2: Scenario parameters

| **Parameters** | **School low risk** | **School high risk** |
|---|---|---|
| $R_{0,S}$: school reproduction number | 1.1 (equal to $R_{0,C}$) | 2.5 |
| $p_s$ : probability of transmission in school | $p_s = p_c$ = 0.0122 (equal to $p_c$, all children-children contacts occur in the community) | $p_s$ =0.0258, $p_c$ =0.0122 |

**Expected percent of index cases with detected forward transmission**

To assess the probability that an index case would have a detected onward transmission event, we developed a statistical model using a Markov process of symptom-based forward tracing that follows a series of steps each with its own probability.

Starting with an infected index case, the first step is to determine whether the index case transmitted, and if so, to how many other individuals. If the index case is transmitted onwards, the next step is whether or not each of the secondary infections arising from it are symptomatic. We assume that if a secondary case is symptomatic, they have a 100% probability of being detected and reported. Of note, this is an optimistic assumption as not all symptomatic contacts are willing to test [11]. If the index case both transmitted onwards and one or more of those transmission is symptomatic and detected, the index case is counted as having a symptom-gated forward transmission (Figure S2). This process is repeated for 1000 index cases to find the probability of detecting a forward transmission starting from an index case for different levels of overdispersion and symptomatic rates.

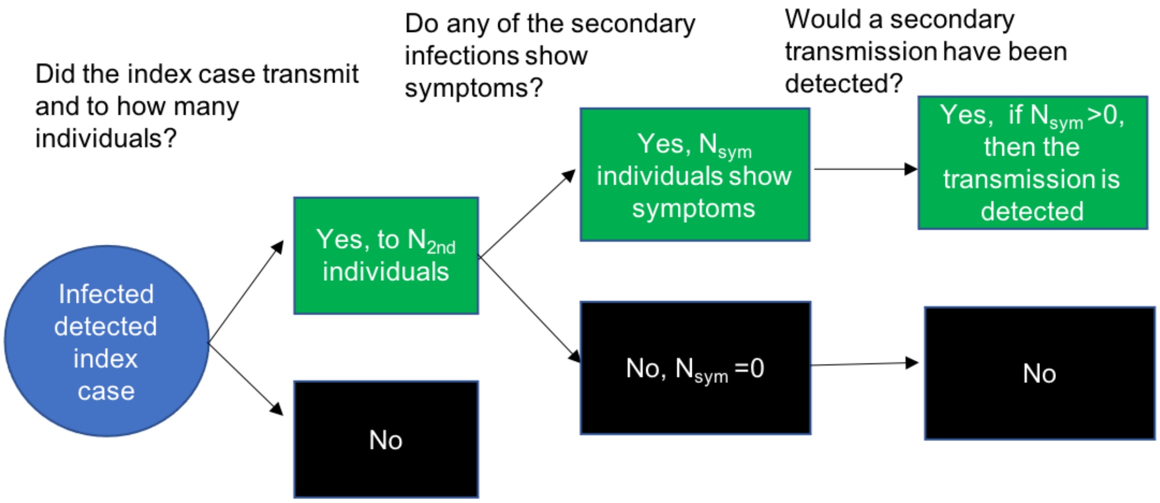


**Figure S2. Schematic diagram of Markov process to model evaluation of efficacy of forward contact-tracing.** We use a Markov process conditioned on having a symptomatic index case, which first assesses whether the index case transmitted onwards, with the number of individuals each index case transmits to following a negative binomial distribution with dispersion parameter $k$ and mean of $R_0$. If the index case transmitted to other individuals, we ask how many of them would be expected to show symptoms, with the number symptomatic following a binomial distribution with a $p_{sym}$ chance of being symptomatic. If an index case is expected to have infected any individuals who would go on to show symptoms, they are reported as having driven a secondary transmission.

The distribution of the number of secondary infections stemming from each index case is dictated by a dispersion parameter, $k$. SARS-CoV-2 is unusual compared to other respiratory diseases like flu in that it has this phenomena called overdispersion, which means that a high proportion of infected individuals transmit to very few individuals, while a small proportion of individuals infect a high number of individuals. The overdispersion parameter *k* of COVID-19 is estimated to have a median value of $k = 0.1$, with a 95% credible interval between 0.05 and 0.2 [12].A higher dispersion factor (*k*) corresponds to a more even distribution of cases. At $k = 0.1$, 20% of infected individuals account for about 80% of secondary transmissions, consistent with observed "super-spreader" phenomena of COVID-19. Crucially, this overdispersion means that it is expected that most people do not actually transmit to anyone else, making negative results about onward transmission difficult to draw conclusions from.

Using previously reported estimates of individual level variation of COVID-19 transmission dynamics [12], we assumed that the number of individuals that an individual went on to infect, $N_{2nd}$ (for number of secondary transmission), follows a negative binomial distribution with an overdispersion parameter *k* and an average number of secondary transmissions, given by the reproduction number, $R_0$.

$$N_{2nd} \sim NegBinom(R_0, k)$$

We assume an $R_0$ of 2.5 to be consistent with our transmission model simulations.
For each index case, the number of secondary transmissions, $N_{2nd}$, is assumed to have a probability $p_{sym}$ of later developing symptoms, such that the number of symptomatic secondary cases corresponding to an index case, $N_{sym}$, is a random variable that follows a binomial distribution.

$$N_{\text{sym}} \sim Binom(N_{2nd}, p_{\text{sym}})$$

This sampling process was repeated 1000 times for the 191 index cases to simulate the forward-tracing process of cases identified in the school in Wood County Wisconsin [6]. To identify the probability that an index case has any detected secondary cases, $p_{detected}$, we count the number of index cases for which $N_{sym}$ is greater than one relative to the total number of index cases.

$$p_{\text{detected}} = \frac{\sum N_{\text{sym}} > 0}{N_{\text{index cases}}}$$

In Figure 2B, we display results for the k = 0.1, 0.05, and 0.2 corresponding to the median and lower and upper bound estimates, respectively, of dispersion for SARS-CoV-2 [12]. The percent symptomatic was varied from 0 to 100%, with dashed and solid lines indicating the symptomatic rates of children and adults respectively [10]. The results shown in Figure 2 indicate that, as overdispersion decreases (high *k*), the percent of index cases detected to transmit will be dictated purely by the percent symptomatic (Figure 2B, blue line). In contrast, when overdispersion is high (low *k*), the percent of index cases detected to transmit is expected to decline for all symptomatic rates, as fewer individuals go on to transmit to others (Figure 2B, green line), and the superspreader that does transmit will transmit to many, making it likely that one of them is detected. At the median estimated level of overdispersion of $k=0.1$, the percent of index cases that transmit is expected to be low (28% [21.5%-34.0%])and relying on symptomatic detections only will render the observed rate even lower, making forward transmission events an ineffective means of quantifying the degree of transmission happening in an environment.

Our results show that forward contact-tracing for SARS-CoV-2 as currently implemented by schools is intrinsically unlikely to detect onwards transmission from a given index case, even if transmission were in fact occurring at a high level. It has been previously shown that forward contact-tracing for SARS-CoV-2 is minimally effective at mitigating transmission [13], and our results demonstrate that it is also minimally effective at estimating the degree of transmission.

### Sensitivity analysis on interactions between adults and children

In the main text, we examined the scenarios in which adults and children's contacts remained constant but child-child contacts had a higher probability of leading to a transmission if schools were opened. We were interested in how the true infection rate (cumulative percent infected) and the discrepancy between true infection rates for adults and children varied as a function of these interactions. We performed a sensitivity analysis to evaluate the effect of increasing the adult-to-child and child-to-adult contacts by a certain factor on the total true infection rate (in both the adult and child populations combined) and the ratio of child to adult true infection rates.

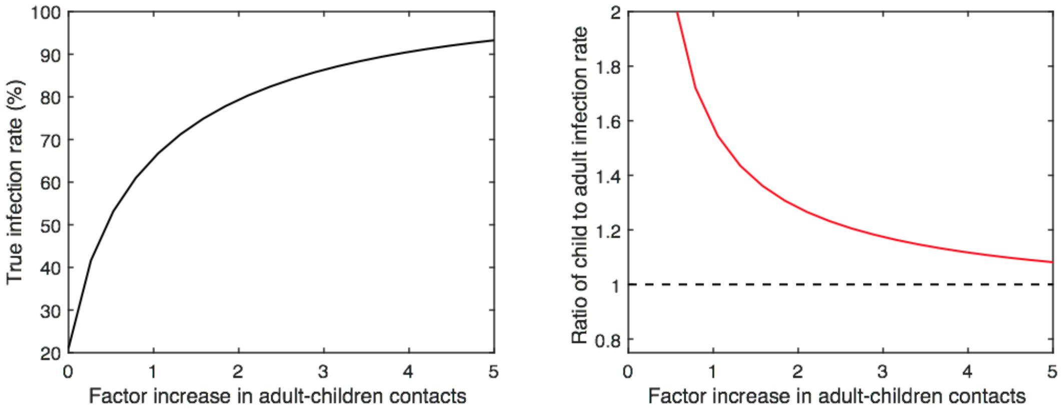


**Figure S3. As school and community interaction increases, the true infection rate goes up, while the ratio of detected case rates looks more similar.** The left figure displays the true infection rate of the adult and child populations combined (cumulative percent infected) as a function of the relative increase in contacts between adults and children (where 1 is the baseline). The right figure displays the ratio of true infection rates in children vs adults as a function of the factor increase in adult-children contacts. Both are for the scenario where within-school $R_{0,S}$ = 2.5 and between-adult $R_{0,C}$ = 1.1.

The sensitivity analysis demonstrates that, as expected, as more mixing between the (faster-spreading) children and the (slower-spreading) adults takes place, more individuals become infected. As a result of the increased potential for cross-over infections from children to adults, greater mixing results in a rapid equilibration of infection rates between school and community. Thus, increased mixing leads at once to a greater total number of infections within the population as a whole, as well as a smaller difference in infection rates between the two populations. Comparing the two rates side by side highlights how comparing infection rates among schools and communities is not an accurate way of assessing the extent of school amplification. If detected cases are estimated by symptom-gated testing, then the higher asymptomatic rates in children can be expected to compound this effect, making it appear that detected case rates in schools are lower than those of the community, even if - as in this scenario- schools are actually driving transmission in the population as a whole.

### References

1. Ismail SA, Saliba V, Bernal JL, Ramsay ME, Ladhani SN. SARS-CoV-2 infection and transmission in educational settings: a prospective, cross-sectional analysis of infection clusters and outbreaks in England. The Lancet Infectious Diseases. 2021. pp. 344–353. doi:10.1016/s1473-3099(20)30882-3

2. Gandini S, Rainisio M, Iannuzzo ML, Bellerba F, Cecconi F, Scorrano L. No evidence of association between schools and SARS-CoV-2 second wave in Italy. doi:10.1101/2020.12.16.20248134

3. Stein-Zamir C, Abramson N, Shoob H, Libal E, Bitan M, Cardash T, et al. A large COVID-19 outbreak in a high school 10 days after schools' reopening, Israel, May 2020. Eurosurveillance. 2020. doi:10.2807/1560-7917.es.2020.25.29.2001352

4. Yung CF, Kam K-Q, Nadua KD, Chong CY, Tan NWH, Li J, et al. Novel Coronavirus 2019 Transmission Risk in Educational Settings. Clin Infect Dis. 2021;72: 1055–1058.

5. Prem K, Cook AR, Jit M. Projecting social contact matrices in 152 countries using contact surveys and demographic data. PLoS Comput Biol. 2017;13: e1005697.

6. Falk A, Benda A, Falk P, Steffen S, Wallace Z, Høeg TB. COVID-19 Cases and Transmission in 17 K-12 Schools - Wood County, Wisconsin, August 31-November 29, 2020. MMWR Morb Mortal Wkly Rep. 2021;70: 136–140.

7. Zimmerman KO, Akinboyo IC, Brookhart MA, Boutzoukas AE, McGann K, Smith MJ, et al. Incidence and Secondary Transmission of SARS-CoV-2 Infections in Schools. Pediatrics. 2021. doi:10.1542/peds.2020-048090

8. Backer JA, Klinkenberg D, Wallinga J. Incubation period of 2019 novel coronavirus (2019-nCoV) infections among travellers from Wuhan, China, 20-28 January 2020. Euro Surveill. 2020;25. doi:10.2807/1560-7917.ES.2020.25.5.2000062

9. He X, Lau EHY, Wu P, Deng X, Wang J, Hao X, et al. Temporal dynamics in viral shedding and transmissibility of COVID-19. Nat Med. 2020;26: 672–675.

10. Davies NG, Klepac P, Liu Y, Prem K, Jit M, CMMID COVID-19 working group, et al. Age-dependent effects in the transmission and control of COVID-19 epidemics. Nat Med. 2020;26: 1205–1211.

11. Doyle T, Kendrick K, Troelstrup T, Gumke M, Edwards J, Chapman S, et al. COVID-19 in Primary and Secondary School Settings During the First Semester of School Reopening - Florida, August-December 2020. MMWR Morb Mortal Wkly Rep. 2021;70: 437–441.

12. Endo A, Centre for the Mathematical Modelling of Infectious Diseases COVID-19 Working Group, Abbott S, Kucharski AJ, Funk S. Estimating the overdispersion in COVID-19 transmission using outbreak sizes outside China. Wellcome Open Res. 2020;5: 67.

13. Endo A, Centre for the Mathematical Modelling of Infectious Diseases COVID-19 Working Group, Leclerc QJ, Knight GM, Medley GF, Atkins KE, et al. Implication of backward contact tracing in the presence of overdispersed transmission in COVID-19 outbreaks. Wellcome Open Res. 2020;5: 239.
